# Causal Effect of Multi-cohort Circulating Proteome on the Risk of Aortic Aneurysm: A Mendelian Randomization Study

**DOI:** 10.1101/2024.11.17.24317384

**Authors:** Yuan Zheng, Lin Qin, Jiayu Ji, Huanqi Mo, Kan Wang

## Abstract

**Background:** The pathogenesis of aortic aneurysm (AA) remains unclear, and there are no effective therapeutic drugs or targets. Circulating plasma proteins are considered biomarkers of AA and potential therapeutic targets for AA. This study aimed to systematically evaluate the causal effects of plasma proteins on AA using a multi-cohort Mendelian randomization (MR) approach.

**Methods:** Protein quantitative trait loci (pQTLs) was obtained from 9 published proteome genome-wide association studies (GWAS) and AA GWAS data from the FinnGen cohort. Independent pQTLs were selected as instrumental variables (IVs). Two-sample MR analysis was performed using inverse-variance weighted, MR-Egger regression, weighted median, weighted mode, and simple mode methods. Heterogeneity and pleiotropy were assessed using Cochran’s Q test, I² statistic, MR-Egger intercept, MR-PRESSO, and Leave-one-out analysis. Steiger filtering was used to test the causal direction. Colocalization analysis and pQTL-eQTL overlap assessment were conducted to validate the findings. Pathway enrichment and drug target analyses were performed to explore the biological and clinical implications of the MR results.

**Results:** A total of 8,285 pQTLs for 4,421 proteins were retained as IVs. Using cis-pQTLs for IVs, MR analysis identified 154 proteins causally associated with TAA (76 protective factors and 78 risk factors) and 211 proteins with AAA (112 protective factors and 99 risk factors). Using cis-pQTLs+trans-pQTLs for IVs, MR analysis identified 236 proteins causally associated with TAA (113 protective factors and 123 risk factors) and 309 proteins with AAA (143 protective factors and 166 risk factors). The MR results showed no significant heterogeneity or pleiotropy. Steiger filtering confirmed the causal direction from circulating proteins to AA. Colocalization analysis found evidence of shared causal variants between multiple proteins and AA. The majority of AA-associated proteins had pQTLs overlapping with blood eQTLs or proxy eQTLs. Pathway enrichment analysis revealed that these proteins were involved in stress response, immune regulation, cytokine-cytokine receptor interaction, metabolic processes and so on. Nearly two-thirds of the causally related proteins were classified as druggable or potentially druggable targets.

**Conclusions:** This study identified a large number of potentially novel pathogenic proteins and therapeutic targets for AA, providing important references for elucidating the molecular pathogenesis of AA and advancing drug development.

## Introduction

Aortic aneurysm (AA) is a life-threatening disease, mainly including abdominal aortic aneurysm (AAA) and thoracic aortic aneurysm (TAA). It is estimated that the number of AA-related deaths worldwide is around 200,000 per year[1–3]. At present, the mechanism of AA formation and progression is not fully understood, and surgical treatment (including open surgery and endovascular repair) is still the most effective treatment for AA patients, and there is no effective drug prevention or treatment of AA[3]. Due to the high mortality, high incidence of complications and high cost of surgical treatment, there is an urgent need to further study the pathogenesis of AA and find effective therapeutic drugs or targets to alleviate the burden of the disease on society[3, 4].

The mechanisms underlying the pathogenesis of AA include the VSMC phenotype switch, apoptosis, oxidative stress, the role of shear stress, the release of various cytokines, the involvement of reactive oxygen species (ROS) leading to aortic cell injury and so on[3]. Recent studies have found that changes in circulating plasma proteins are closely related to the development and treatment of AA[5]. As the basic functional unit of the human body, proteins are the largest class of drug targets[6]. Proteomic studies have found that specific proteins can serve as biomarkers for AA and may be related to the diagnosis, development, treatment, and pathogenesis of AA[7, 8]. In recent years, researchers have increasingly utilized proteomics methods to study body fluids, aiming to uncover disease mechanisms related to the cardiovascular system and improve the diagnosis, prognosis, and monitoring of cardiovascular diseases (CVD)[9]. However, research on the relationship between circulating plasma proteins and the pathogenesis of AA is insufficient, and most existing studies are observational and focus on the correlation between a single or several circulating proteins and AA, which also has limitations in inferring causality[10]. Therefore, it is necessary to find more effective analytical methods to explore the causal relationship between AA and circulating proteins.

Mendelian randomization (MR) is a widely used method of causal inference which can better control for confounding factors, minimize reverse causality effects, reduce bias risk and enhance the effectiveness of causal inference[11, 12]. MR studies use genetic variants associated with specific protein levels as instrumental variables (IVs), simulating a process similar to a randomized controlled trial based on the principle of random allocation of alleles in Mendel’s laws. Genetic variation is not influenced by traditional risk factors and precedes disease occurrence without being altered by disease outcomes, thus largely avoiding confounding bias and reverse causality in traditional epidemiological studies. In addition, the flourishing development of genome-wide association study (GWAS) data also provides a good foundation for the implementation of MR studies.

Based on GWAS data, this study aims to explore the causal association between circulating plasma proteins and AA through MR analysis.

## Methods

### Data Sources

pQTLs (Protein Quantitative Trait Loci) were derived from 9 published plasma proteome GWASs, genetic data associated with TAA from the FinnGen cohort (3880 cases and 381977 controls) and AAA from the FinnGen cohort (3869 cases and 381977 controls)[13]. The website(https://www.finngen.fi/en) was accessed on April 15, 2024(Table 1). The original studies have obtained informed consent from the study subjects, so this part of the study does not involve the need for ethics committee approval.

**Table 1.**
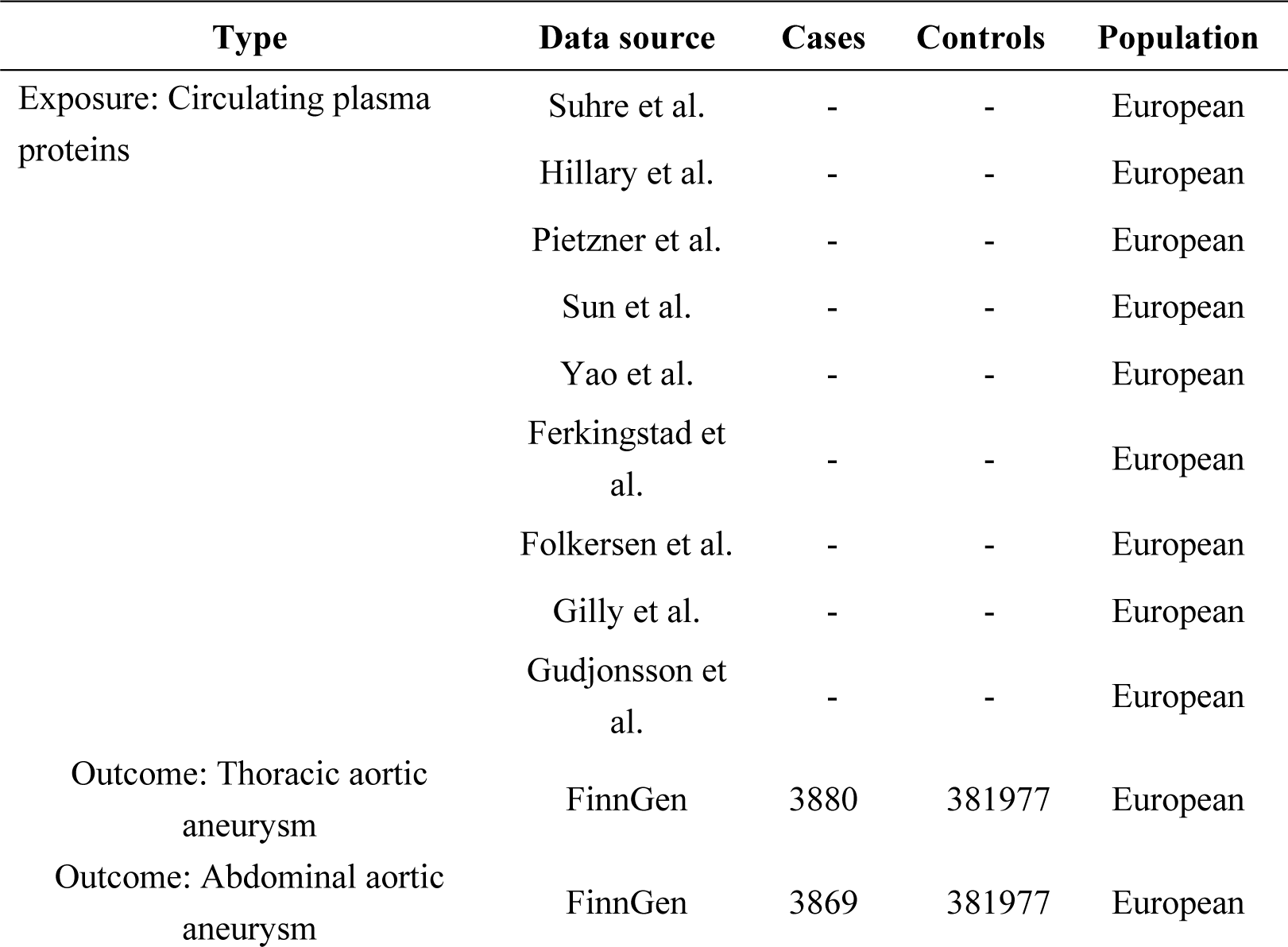
Brief information on pQTLs and GWAS databases in MR studies

| Type | Data source | Cases | Controls | Population |
| --- | --- | --- | --- | --- |
| Exposure: Circulating plasma proteins | Suhre et al. | - | - | European |
|  | Hillary et al. | - | - | European |
|  | Pietzner et al. | - | - | European |
|  | Sun et al. | - | - | European |
|  | Yao et al. | - | - | European |
|  | Ferkingstad et al. | - | - | European |
|  | Folkersen et al. | - | - | European |
|  | Gilly et al. | - | - | European |
|  | Gudjonsson et al. | - | - | European |
| Outcome: Thoracic aortic aneurysm | FinnGen | 3880 | 381977 | European |
| Outcome: Abdominal aortic aneurysm | FinnGen | 3869 | 381977 | European |

### IVs screening for pQTLs associated with AA

MR IVs for circulating proteins were constructed based on 9 proteomic GWAS data (screening criteria: sample size > 500, proteins measured > 50). IVs were divided into cis-pQTLs and trans-pQTLs. Cis-pQTLs were defined as pQTLs within a 500kb window of the corresponding protein-coding sequence, while trans-pQTLs were defined as pQTLs outside the 500kb window of the protein-coding gene.

① Single nucleotide polymorphisms (SNPs) associated with any protein were selected based on the P-value threshold recommended in the respective studies[14].

②SNPs located within the major histocompatibility complex (MHC) region (chr6: 26Mb to 34Mb) were removed because the linkage disequilibrium (LD) patterns of SNPs in the MHC region are complex.

③ LD cluster analysis was performed with a threshold of r^2^>0.01 and upstream/downstream distance <5000kb to determine independent pQTLs for each protein.

④ Instrumental variables associated with five or more proteins were excluded because these instrumental variables are highly pleiotropic.

⑤Palindromic SNPs were excluded, and SNPs directly associated with the outcome variable (P<1×10^-5^) were excluded.

The analysis framework of this study is shown in Figure 1.

**Figure 1.**
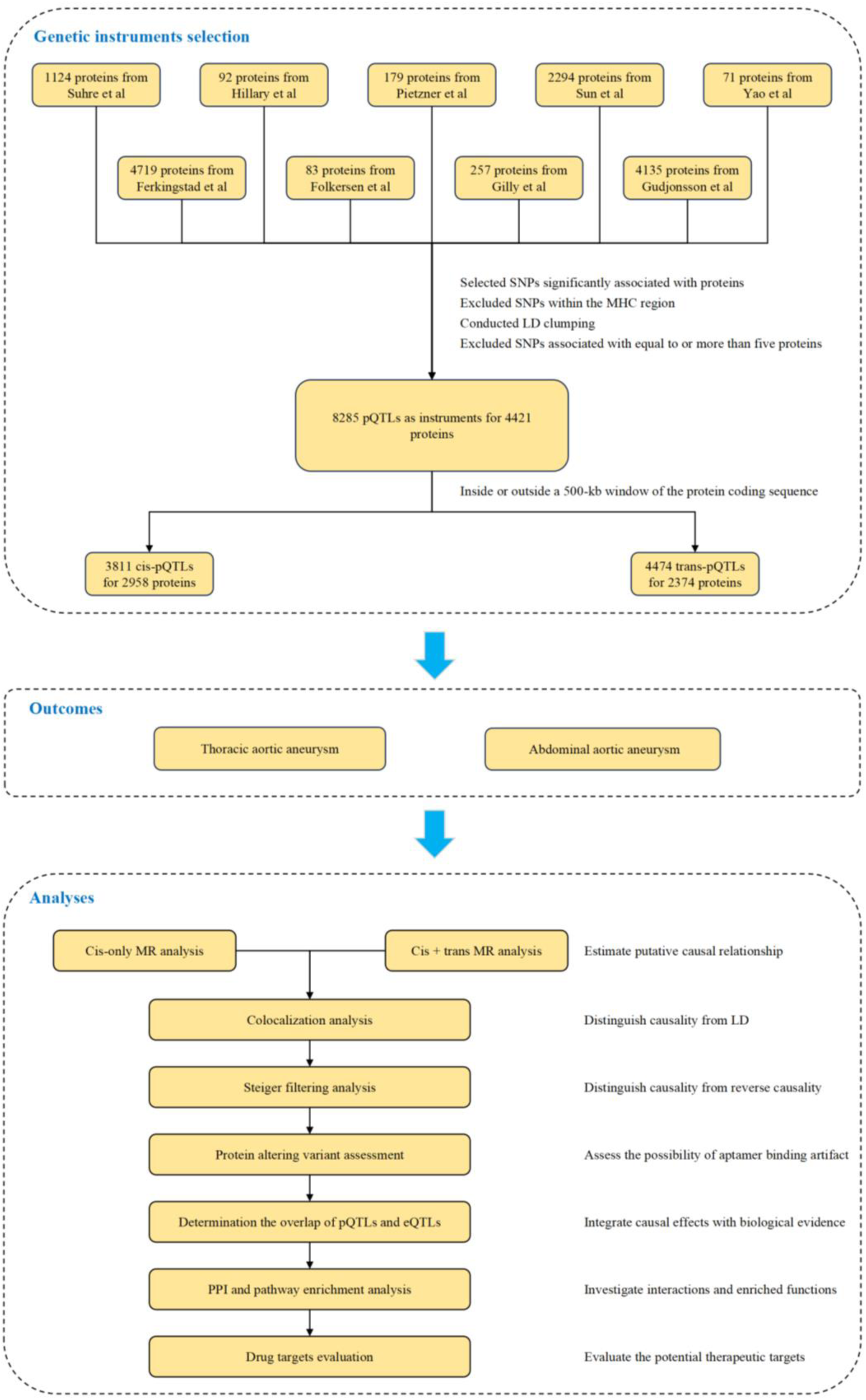
Flowchart of the MR analysis framework for evaluating the effect of multi-cohort plasma proteome on AA.

### MR analysis estimates the effect of circulating proteins on AA

5 regression models, MR-Egger regression, random-effects inverse variance weighted (IVW), weighted median, weighted mode, and simple mode, were used to assess the potential causal relationship between exposure factors (circulating proteins) and outcome (TAA, AAA) risk using pQTLs as IVs in a two-sample MR analysis. Random-effects IVW was the main causal analysis method, and other methods were supplementary analysis methods. When SNPs≤3, the effect of individual SNPs on the outcome was estimated using the Wald ratio method, and the rest were estimated using fixed-effects IVW; when more than 3 SNPs were included, random-effects IVW was used. For causal effect estimation based on multiple SNPs as IVs, IVW weights each locus’s causal effect estimate by the inverse of its variance (R^2), sums the weighted estimates for each locus, and the final estimate is the causal effect estimate of the IVW method. Cochran’s Q test was used to determine SNP heterogeneity, with *P*<0.05 indicating heterogeneity in the results. I^2^ (I-squared) is another measure of heterogeneity, representing the proportion of total variation caused by heterogeneity. I² ranges from 0% to 100%, with I² greater than 50% indicating some heterogeneity in IVW results. The formula is 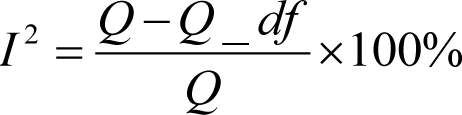 MR-Egger’s intercept and MR-PRESSO are used for pleiotropy analysis, and Leave-one-out for sensitivity analysis. MR-Egger regression intercept being equal to zero and not statistically significant (P > 0.05), along with MR-PRESSO P>0.05 indicated no pleiotropy of SNPs. Leave-one-out analysis of SNPs is conducted by progressively removing each SNP, reanalyzing with the remaining SNPs, and observing the impact of each SNP on the analysis results. The above methods were implemented using the Two Sample MR package in R software version 4.1.0, with a significance level of α = 0.05. To effectively control the false discovery rate, P-values were corrected using the Benjamini-Hochberg method. When SNPs ≤ 3, MR-PRESSO cannot be used for pleiotropy analysis.

### Testing causal direction using Steiger filtering analysis

The Steiger filtering test is a statistical method used to test the consistency of the causal relationship direction of genotypes to intermediate variables and final outcomes. In the MR Steiger results, the variance of the outcome being less than the variance of the circulating plasma protein was judged as “TRUE”, indicating that the causal relationship was consistent with the expected direction, while a “FALSE” result indicated that the causal relationship was opposite to the expected direction.

### Colocalization analysis

SNPs within ±500kb of the gene’s cis-acting region were extracted using the R package “coloc” for colocalization analysis. Colocalization analysis is commonly used to identify whether two phenotypes are driven by the same causal variant in a given region, thus strengthening the evidence of association between the two phenotypes. In a given region, colocalization analysis makes the prior assumption that each of the two traits has at most 1 truly causal variant in that region, resulting in five mutually exclusive model assumptions (H0-H4), which are all possible association scenarios under the prior assumption.

H0: Phenotype 1 (GWAS) and phenotype 2 (QTL or GWAS) are not significantly associated with any SNP loci in a genomic region.

H1/H2: Phenotype 1 or phenotype 2 is significantly associated with SNP loci in a genomic region.

H3: Phenotypes 1 and 2 are significantly associated with SNP loci in a genomic region, but are driven by different causal variant loci.

H4: Phenotypes 1 and 2 are significantly associated with SNP loci in a genomic region and are driven by the same causal variant loci.

During the colocalization analysis, posterior probabilities (PP.H0-PP.H4) are generated for each of the above models, with the sum of the five model posterior probabilities being 1. The higher the posterior probability of a model, the more likely the corresponding model assumption is to hold given the data. Generally, we prefer the H4 assumption to hold, because the H4 model assumption indicates that the two traits are driven by the same causal variant. Generally, when PP.H4>0.75, we consider the H4 model assumption to hold.

### Assessing the degree of overlap between pQTLs and eQTLs

Genetic variation can quantitatively influence transcript and protein levels. Existing eQTLs corresponding to pQTLs prove that their genetic effects on protein expression are achieved by regulating the corresponding mRNA transcription, which can improve the biological interpretability of pQTLs. To explore the potential mechanism of pQTLs on circulating protein levels, we investigated the degree of overlap between pQTLs and eQTLs through direct genetic lookup. For pQTLs with MR evidence, blood eQTL data provided by the IEU OpenGWAS project (https://gwas.mrcieu.ac.uk/) were used to detect whether the variants themselves and proxies (r^2^≥0.8) had corresponding eQTLs.

### Pathway enrichment analysis of proteins with causal effects in MR analysis

To investigate the relationships among proteins with causal effects in MR analysis, gene ontology (GO) and Kyoto Encyclopedia of Genes and Genomes (KEGG) pathway enrichment analyses were performed using the online web tool (https://biit.cs.ut.ee/gprofiler/gost) to explore potential enriched pathways associated with these proteins.

### Assessment of drug targets for proteins exhibiting causal effects in MR **analysis**

To assess whether proteins with causal effects in MR analysis overlap with genes from the druggable genome, we compared proteins with causal effects in MR analysis with the list of druggable genes by Finan[15]. Finan and colleagues systematically identified 4,479 genes as the druggable or potentially druggable genome and classified them into three tiers based on their position in the drug development pipeline. Tier 1 (1,427 genes) includes targets of approved small molecule and biotherapeutic drugs as well as clinical-phase drug candidates; Tier 2 consists of 682 genes encoding targets with known bioactive drug-like small molecule binding partners and proteins with ≥ 50% identity (over 75% of the sequence) to approved drug targets, and Tier 3 includes 2,370 genes encoding secreted or extracellular proteins, proteins distantly related to approved drug targets, and members of key druggable gene families not already included in Tier 1 or 2. This tier is further subdivided, with priority given to genes close (±50kb) to GWAS SNPs and with extracellular location (Tier 3A), and the remaining genes assigned to Tier 3B. In our analysis, we focused on the following information for proteins with causal effects in MR analysis: priority of druggable genes; predicted to be targeted if the protein product of the gene is targeted by a small molecule; predicted to be targeted if the protein product of the gene is targeted by a biotherapy (monoclonal antibody/enzyme or other protein).

In addition, drug target annotation of proteins with causal effects in MR analysis was performed based on the Therapeutic Target Database (http://db.idrblab.net/ttd/). This database provides 4,298 drug targets, including 803 successful drug targets, 1,327 clinical trial targets, 240 clinical trial/patent targets, 1,845 research targets, and 83 discontinued targets. We focused on the type of drug target, the drug used for the target, and the disease targeted by the drug.

## Results

### Screening Results of Instrumental Variables

pQTLs were screened from 9 proteomic GWAS data as genetic IVs. After screening, 8,285 pQTLs for 4,421 proteins (2,518 unique proteins) were retained as tools for MR analysis (Supplemental Table 1). Ivs were further divided into cis-pQTLs and trans-pQTLs, with 3,811 cis-pQTLs used for 2,958 proteins (1,558 unique proteins) and 4,474 trans-pQTLs used for 2,374 proteins (1,763 unique proteins) (Supplemental Table 2-1 and Supplemental Table 2-2). Among the 4,421 proteins with valid instrumental variables, 911 proteins were influenced by both cis-pQTLs and trans-pQTLs, 2,047 proteins were influenced only by cis-pQTLs, and 1,463 proteins were influenced only by trans-pQTLs.

### MR analysis estimates the effect of circulating plasma proteins on AA

We first used cis-pQTLs as genetic tools for MR analysis to systematically evaluate the evidence for causal effects of circulating plasma proteins on AA. The results showed that a total of 365 circulating plasma proteins (256 unique proteins) were causally associated with AA, of which 154 proteins were causally associated with TAA (76 were protective factors and 78 were risk factors for TAA), and 211 proteins were causally associated with AAA (112 were protective factors and 99 were risk factors for AAA) (Figure 2 and Supplemental Table 3). IVW results showed no heterogeneity among cis-pQTLs associated with circulating plasma proteins and AA, MR-Egger result shows that the sum of intercept terms equals 0, no statistically significant difference, MR-PRESSO P-value greater than 0.05, suggesting no horizontal pleiotropy of SNPs and indicating that the MR results of this study are robust.

**Figure 2.**
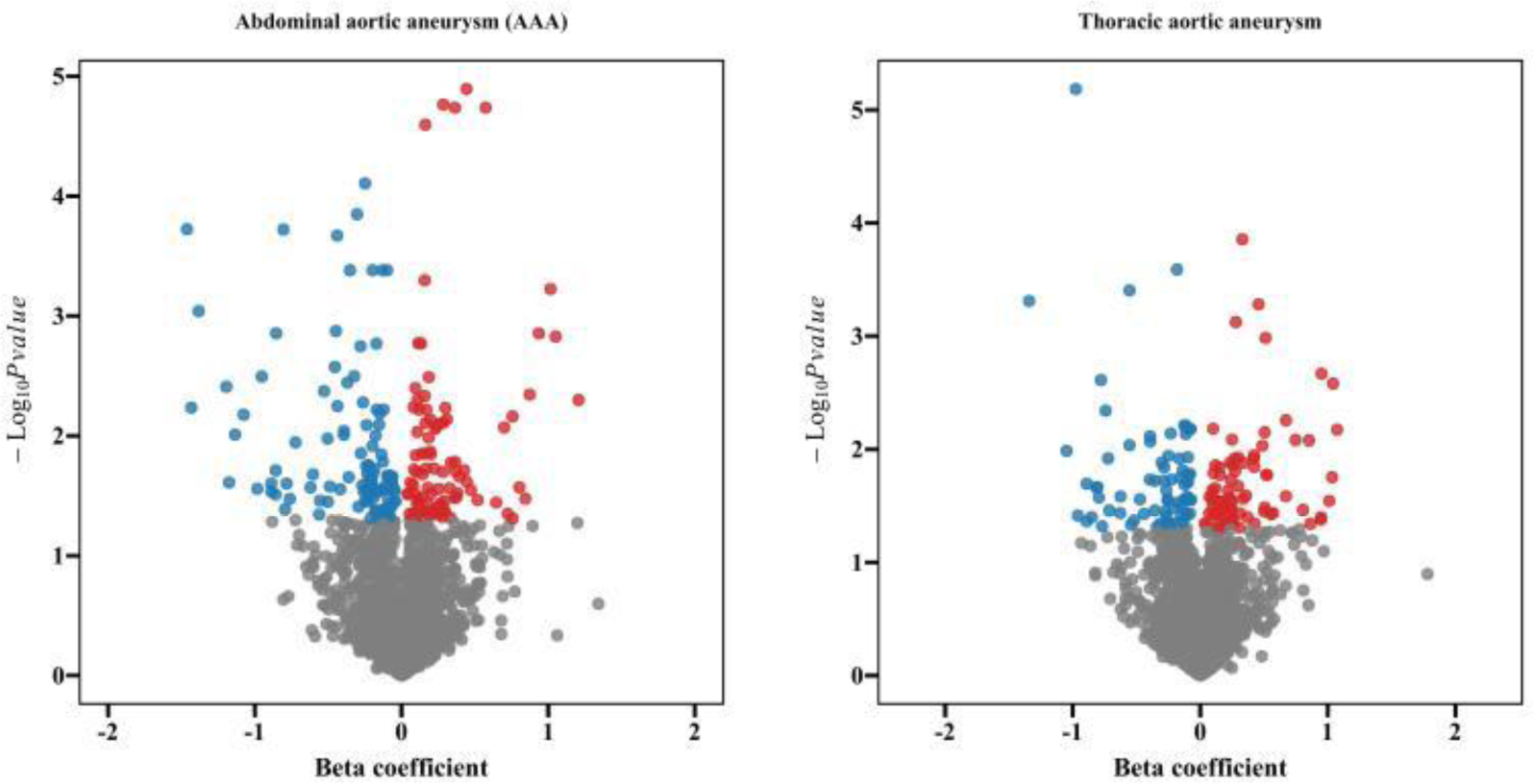
Volcano plot of causal effects of circulating plasma proteins on phenotypes analyzed by cis-pQTLs. Red dots represent circulating plasma proteins significantly associated with the outcome variable and with a beta effect value greater than 0. Blue dots represent circulating plasma proteins significantly associated with the outcome variable and with a beta effect value less than 0. Gray dots represent circulating plasma proteins not significantly associated with the outcome variable.

In MR analysis, adding trans-pQTLs may increase the reliability of the association between proteins and phenotypes. Therefore, we used all (cis + trans) pQTLs as IVs to extend our MR analysis. The results showed that a total of 545 circulating plasma proteins (413 unique proteins) were causally associated with AA, of which 236 proteins were causally associated with TAA (113 were protective factors and 123 were risk factors for TAA), and 309 proteins were causally associated with AAA (143 were protective factors and 166 were risk factors for AAA)(Figure 3 and Supplemental Table 4). Most of these associations did not appear in the cis-pQTLs analysis. Sensitivity analysis showed that all significant associations detected were robust. 134 new circulating plasma proteins were also found to be associated with abdominal AA(Supplemental Table 4).

**Figure 3.**
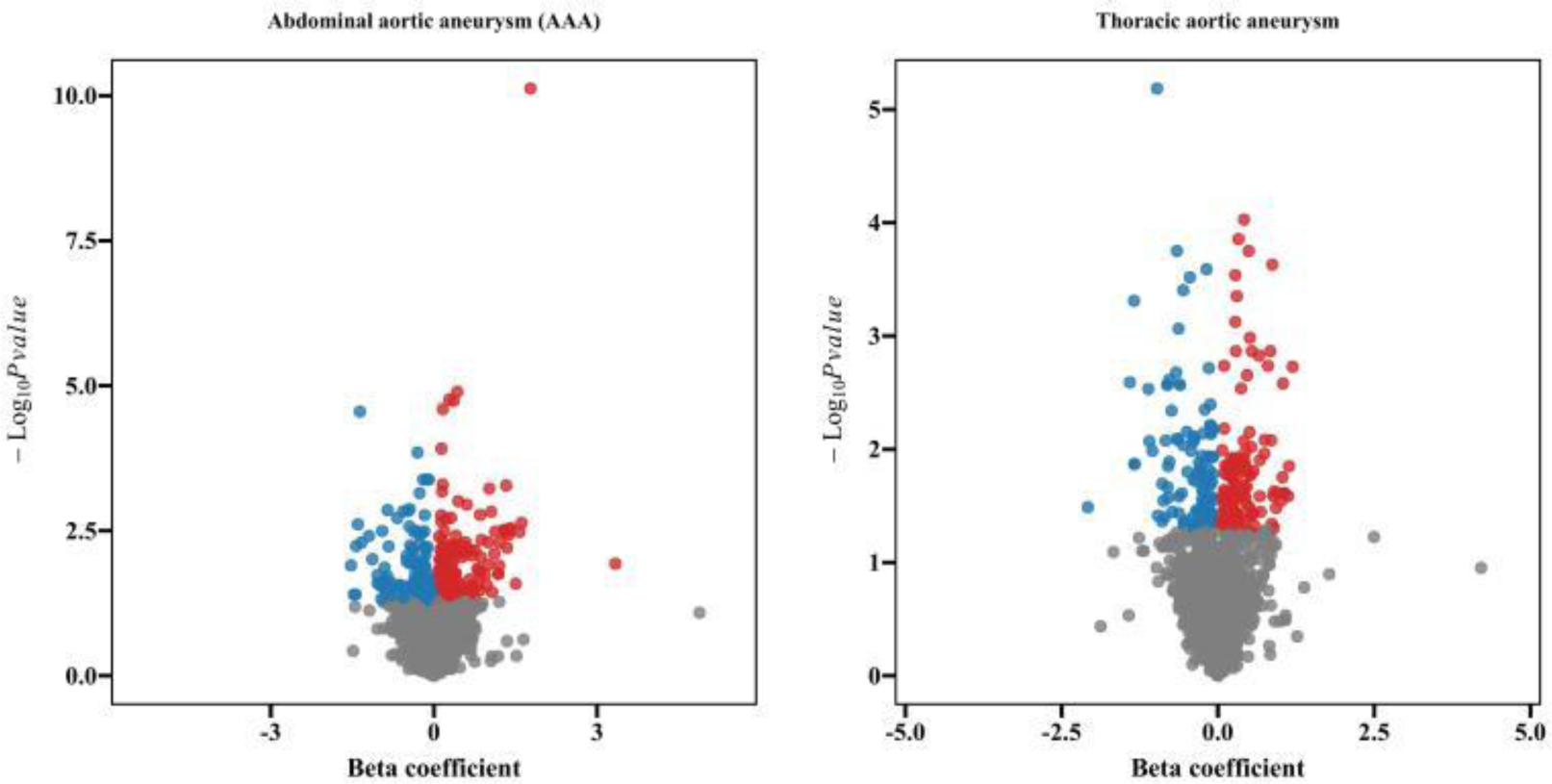
Volcano plot of causal effects of circulating plasma proteins on phenotypes analyzed by cis-pQTLs+trans-pQTLs. Red dots represent circulating plasma proteins significantly associated with the outcome variable and with a beta effect value greater than 0. Blue dots represent circulating plasma proteins significantly associated with the outcome variable and with a beta effect value less than 0. Gray dots represent circulating plasma proteins not significantly associated with the outcome variable.

### Verification of causality and expected direction

Using cis-pQTLs as genetic instrumental variables for MR analysis, the results of the Steiger directionality test show that the direction of circulating plasma proteins in the AA dataset is all “TRUE”, indicating that the causal relationship between circulating plasma proteins and outcomes is consistent with the expected direction.

Using cis-pQTLs+trans-pQTLs as genetic IVs for MR analysis, the results of the Steiger directionality test showed that the directions of circulating plasma proteins in both TAA and AAA dataset were all “TRUE”, indicating that the causal relationship between circulating plasma proteins and outcomes was consistent with the expected direction (Supplemental Table5).

### Colocalization analysis

Using cis-pQTLs as genetic IVs for MR analysis, 365 circulating plasma proteins significantly associated with the outcome variable2 were colocalized with the outcome variable2. Evidence of colocalization existed for pQTLs of 7 circulating plasma proteins with TAA, indicating shared causal variants. Evidence of colocalization for pQTLs of 16 circulating plasma proteins with AAA also suggested shared causal variants. Circulating plasma protein OVCA2 was the most likely causal variant in the colocalization analysis for TAA (PP.H4=0.995). Circulating plasma protein PLAU was the most likely causal variant in the colocalization analysis for AAA (PP.H4=0.993) (Table 2).

**Table 2.** Colocalization analysis of circulating plasma proteins and phenotypes (using cis-pQTLs as genetic IVs for MR analysis)

| Uniprot | Exposure | Source | Outcome | PP.H0 | PP.H1 | PP.H2 | PP.H3 | PP.H4 | SNP | SNP.PP. |
| --- | --- | --- | --- | --- | --- | --- | --- | --- | --- | --- |
| P17405 | SMPD1 | GenAss | AAA | 4.35E-65 | 0.009 | 5.07E-66 | 0 | 0.991 | rs1050239 | 1 |
| P08519 | LPA | GenMap | AAA | 2.60E-69 | 0.014 | 2.05E-70 | 1.42E-04 | 0.986 | rs186696265 | 1 |
| Q92484 | SMPDL3A | GenAtl | AAA | 1.20E-186 | 0.133 | 7.84E-189 | 0 | 0.867 | rs28385609 | 1 |
| O43405 | COCH | GenAss | AAA | 1.72E-61 | 0.066 | 2.44E-63 | 5.39E-06 | 0.934 | rs7159420 | 1 |
| Q99988 | GDF15 | GenAtl | AAA | 6.03E-91 | 0.166 | 3.02E-93 | 0 | 0.834 | rs45543339 | 1 |
| O43405 | COCH | GenAtl | AAA | 6.99E-32 | 0.047 | 1.42E-33 | 0 | 0.953 | rs34907608 | 1 |
| Q8WZ82 | OVCA2 | LarInt | TAA | 1.05E-16 | 0.005 | 2.07E-17 | 0 | 0.995 | rs145234879 | 1 |
| P17405 | SMPD1 | LarInt | AAA | 5.19E-76 | 0.009 | 6.05E-77 | 0 | 0.991 | rs1050239 | 1 |
| P00747 | PLG | GenAss | AAA | 1.47E-21 | 0.073 | 1.87E-23 | 0 | 0.927 | rs2489929 | 1 |
| Q92484 | SMPDL3A | LarInt | AAA | 1.60E-167 | 0.133 | 1.05E-169 | 0 | 0.867 | rs28385609 | 1 |
| Q86TH1 | ADAMTSL2 | LarInt | AAA | 3.76E-32 | 0.081 | 4.26E-34 | 0 | 0.919 | rs11534419 | 1 |
| P00749 | PLAU | GenAtl | AAA | 8.43E-16 | 0.007 | 1.21E-16 | 0 | 0.993 | rs2227551 | 1 |
| Q16674 | MIA | LarInt | AAA | 2.43E-08 | 0.081 | 2.75E-10 | 0 | 0.919 | rs10407351 | 1 |
| Q14392 | LRRC32 | LarInt | TAA | 1.21E-07 | 0.161 | 6.32E-10 | 0 | 0.839 | rs1320646 | 1 |
| Q9BPX1 | HSD17B14 | GenAss | TAA | 1.65E-12 | 0.200 | 6.60E-15 | 0 | 0.800 | rs35299026 | 1 |
| Q9Y251 | HPSE | GenAss | TAA | 3.35E-125 | 0.097 | 3.11E-127 | 0 | 0.903 | rs6535455 | 1 |
| P40763 | STAT3 | LarInt | TAA | 4.62E-72 | 0.149 | 2.64E-74 | 0 | 0.851 | rs4796791 | 1 |
| Q92484 | SMPDL3A | ConGe | AAA | 6.14E-12 | 0.133 | 4.02E-14 | 0 | 0.867 | rs28385609 | 1 |
| P00749 | PLAU | GenAss | AAA | 1.10E-75 | 0.009 | 1.22E-76 | 0 | 0.991 | rs2227564 | 1 |
| P30740 | SERPINB1 | LarInt | AAA | 3.50E-16 | 0.203 | 1.38E-18 | 0 | 0.797 | rs2293772 | 1 |
| Q969Z4 | RELT | GenAss | TAA | 5.85E-53 | 0.204 | 2.28E-55 | 0 | 0.796 | rs73542967 | 1 |
| Q9BWD1 | ACAT2 | LarInt | TAA | 1.53E-194 | 0.063 | 2.27E-196 | 0 | 0.937 | rs25683 | 1 |
| Q92484 | SMPDL3A | GenAss | AAA | 8.37E-190 | 0.133 | 5.48E-192 | 0 | 0.867 | rs28385609 | 1 |

Using cis-pQTLs+trans-pQTLs as genetic IVs for MR analysis, 545circulating plasma proteins significantly associated with the outcome variable2 were colocalized with the outcome variable2. Evidence of colocalization existed for pQTLs of 12 circulating plasma proteins with TAA, indicating shared causal variants. Evidence of colocalization for pQTLs of 18 circulating plasma proteins with AAA also suggested shared causal variants. Circulating plasma protein OVCA2 was the most likely causal variant in the colocalization analysis for TAA (PP.H4=0.995). Circulating plasma protein PLAU and KAZALD1 were the most likely causal variant in the colocalization analysis for AAA (PP.H4=0.993) (Table 3).

**Table 3.**
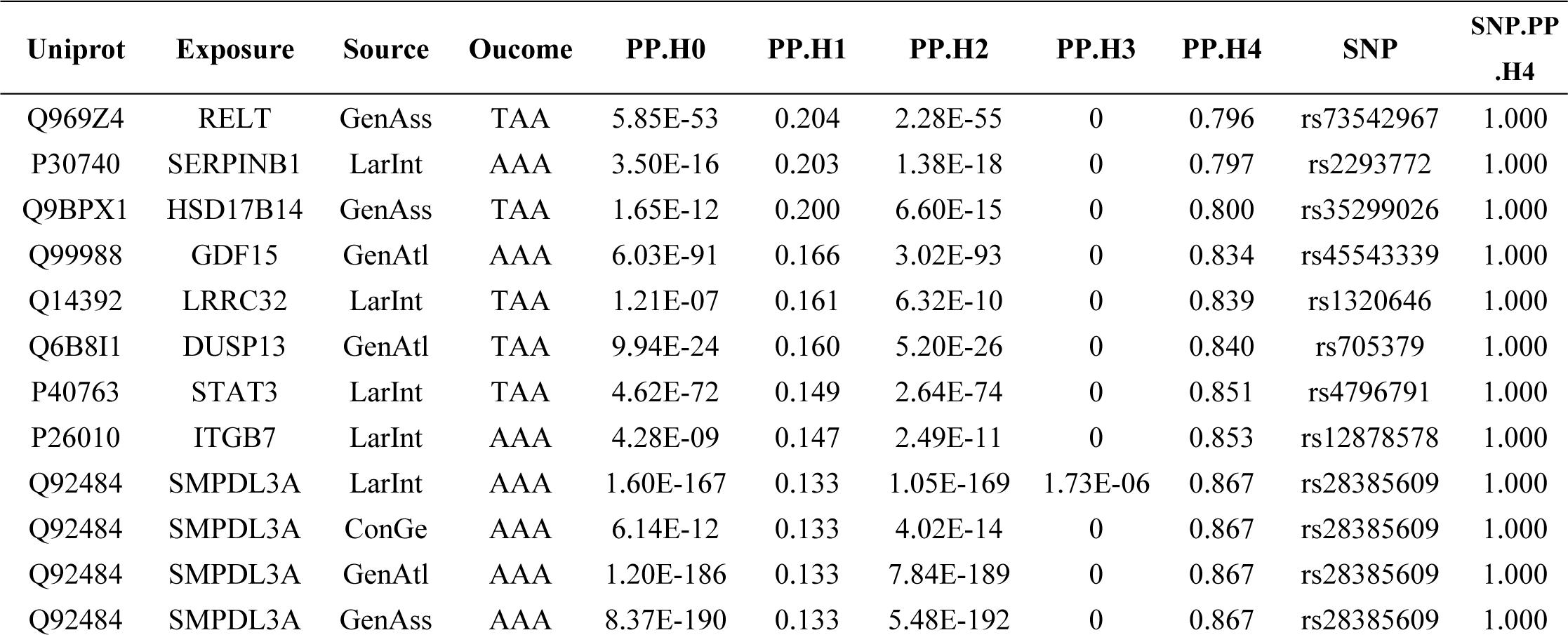
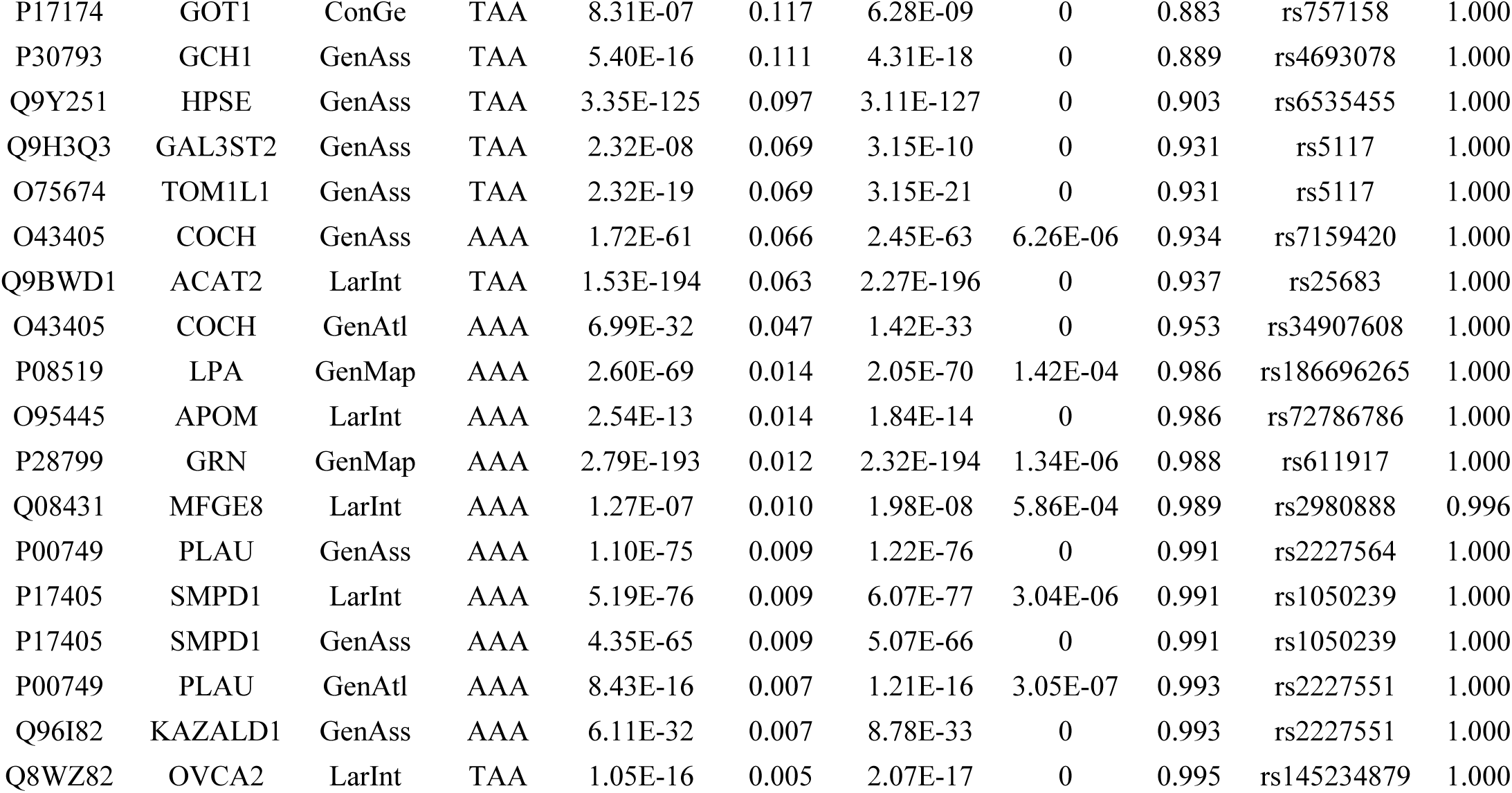
Colocalization analysis of circulating plasma proteins and phenotypes (using cis-pQTLs +trans-pQTLs as genetic IVs for MR analysis)

### Assessing the degree of overlap between pQTLs and eQTLs

To assess the possibility that the association of some pQTLs with circulating plasma protein levels is driven by effects on transcriptional levels rather than other mechanisms, we investigated the degree of overlap between pQTLs and eQTLs. In the results of MR analysis using only cis-pQTLs, the pQTLs of 365 circulating plasma proteins significantly associated with AA(337 unique circulating plasma proteins after de-duplication) were overlapped with eQTLs. 389 cis-pQTLs were used as IVs for proteins with MR evidence, and of these cis-pQTLs, 283 pQTLs overlapped with blood eQTLs. In addition, 0 pQTLs overlapped with blood proxy eQTLs (Supplemental Table 6-1).

In the results of MR analysis using cis-pQTLs+trans-pQTLs, the pQTLs of 545 circulating plasma proteins significantly associated with AA(515 unique circulating plasma proteins after de-duplication) were overlapped with eQTLs. 862 cis-pQTLs were used as IVs for proteins with MR evidence, and of these cis-pQTLs, 326 pQTLs overlapped with blood eQTLs. In addition, 34 pQTLs overlapped with blood proxy eQTLs(Supplemental Table6-2).

### Pathway enrichment analysis of proteins with causal effects in MR analysis

Using cis-pQTLs as genetic tools for MR analysis, 211 circulating plasma proteins significantly associated with AAA were functionally enriched. In GO enrichment analysis, proteins with causal effects in cis-pQTLs MR analysis were enriched in stress response, regulation of multicellular organismal process, and other pathways (Figure 4 A). KEGG pathway analysis revealed that proteins with causal effects in cis-pQTLs MR analysis were significantly enriched in complement and coagulation cascades, IL-17 signaling pathway, and other related pathways (Figure 4 B). 154 circulating plasma proteins significantly associated with TAA were functionally enriched. In GO enrichment analysis, proteins with causal effects in cis-pQTLs MR analysis were enriched in cell chemotaxis, regulation of immune system processes, and other pathways (Figure 4 C). KEGG pathway analysis revealed that proteins with causal effects in cis-pQTLs MR analysis were significantly enriched in cytokine-cytokine receptor interaction and complement and coagulation cascade pathways (Figure 4 D).

**Figure 4.**
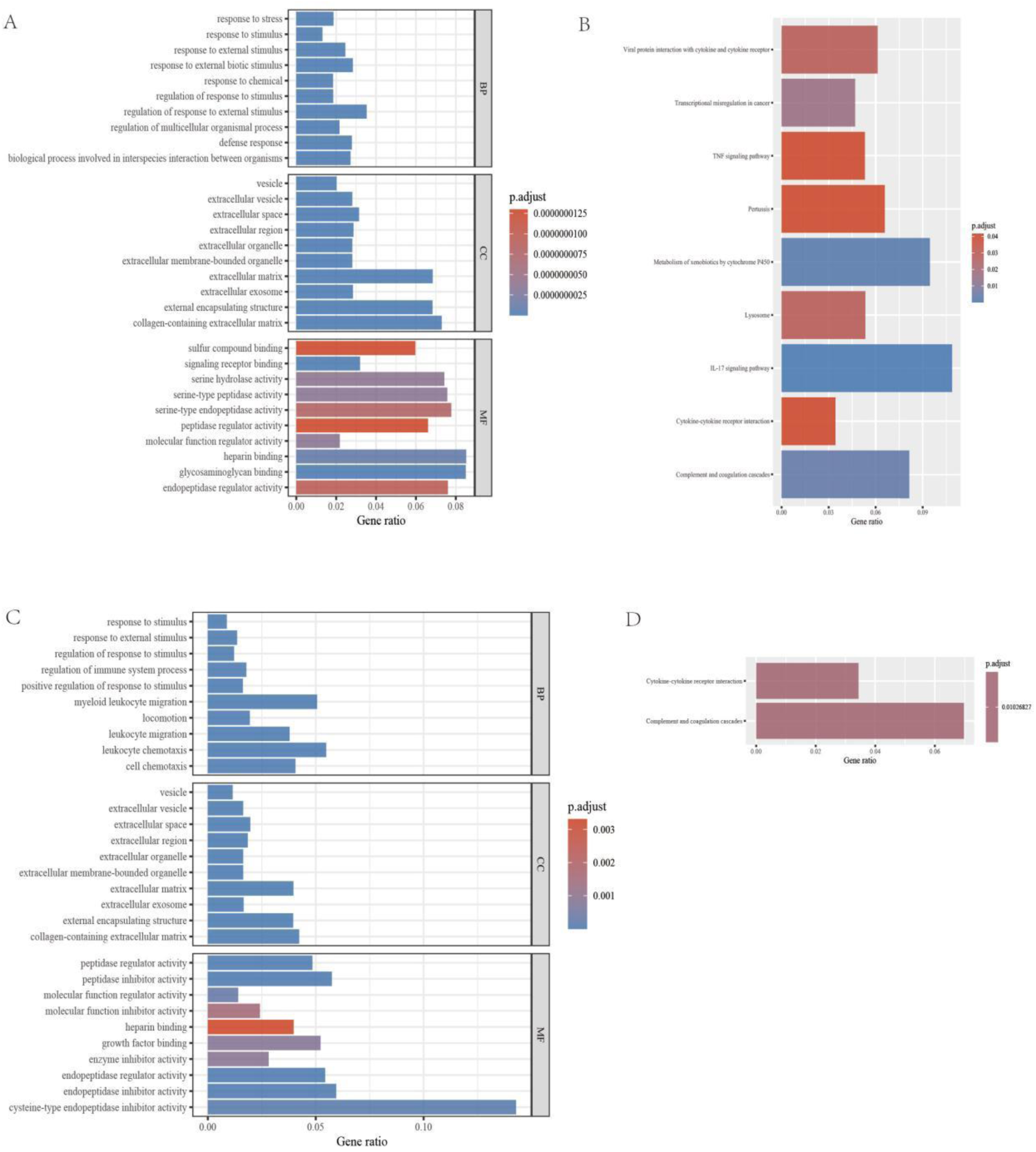
GO and KEGG pathway enrichment analysis of plasma proteins associated with AA (cis-pQTLs MR analysis) (A)GO enrichment analysis of AAA; (B) KEGG pathway analysis of AAA; (C)GO enrichment analysis of TAA; (D) KEGG pathway analysis of TAA.

Using cis-pQTLs+trans-pQTLs as genetic tools for MR analysis, 309 circulating plasma proteins significantly associated with AAA were functionally enriched. In GO enrichment analysis, proteins with causal effects in cis-pQTLs+trans-pQTLs analysis were enriched in response to stimulus, organonitrogen compound metabolic process, and other pathways (Figure 5 A). KEGG pathway analysis revealed no significant enrichment of proteins with causal effects in cis-pQTLs+trans-pQTLs MR analysis. 236 circulating plasma proteins significantly associated with TAA were functionally enriched. In GO enrichment analysis, several pathways related to thoracic AA biology were enriched. Proteins with causal effects in cis-pQTLs+trans-pQTLs MR analysis were enriched in response to stimulus, regulation of multicellular organismal process, cell adhesion, and other pathways (Figure 5 B). KEGG pathway analysis revealed that proteins with causal effects in cis-pQTLs+trans-pQTLs MR analysis were significantly enriched in metabolic pathways, cytokine-cytokine receptor interaction, and other related pathways (Figure 5 C).

**Figure 5.**
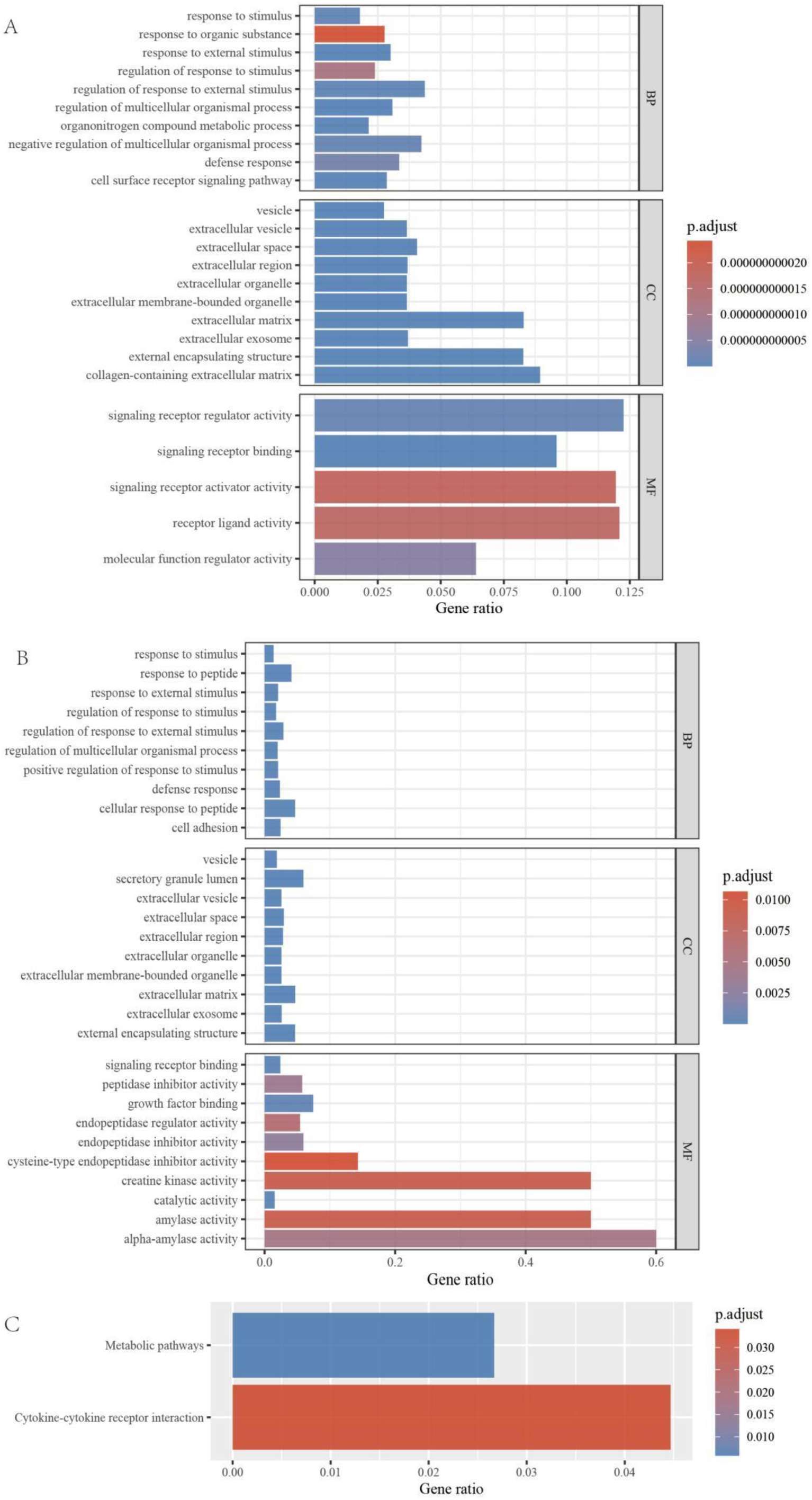
GO and KEGG pathway enrichment analysis of plasma proteins associated with AA (cis-pQTLs+trans-pQTLs MR analysis) (A)GO enrichment analysis of AAA; (B)GO enrichment analysis of TAA; (C) KEGG pathway analysis of TAA.

### Assessing drug targets for proteins with causal effects in MR analysis

A total of 450 circulating plasma proteins were significantly associated with AA in the results of cis-pQTLs and cis-pQTLs+trans-pQTLs analyses. Among these 450 proteins, 270 had druggable targets, of which 70 were Tier 1, 30 were Tier 2, 90 were Tier 3A, and 80 were Tier 3B. Using the Therapeutic Target Database, a total of 123 proteins were found to be targets of existing or potential drugs, including 29 successful drug targets, 49 clinical trial targets, 1 preclinical target, 4 patent record targets, 39 literature reported targets, and 1 discontinued target(Supplemental Table 7).

## Discussion

This study systematically evaluated the causal relationship between circulating protein levels and AA at the proteome-wide level using large-scale plasma proteomic GWAS data and MR analysis framework. The results identified a large number of potentially novel pathogenic proteins and therapeutic targets for AA, providing important references for elucidating the molecular pathogenesis of AA and advancing drug development. MR Analyses using cis-pQTLs, cis-pQTLs+trans-pQTLs as IVs identified hundreds of circulating plasma proteins that were associated with AA, and that the causal direction between these circulating proteins and AA was consistent with expectations and may be influenced by the same underlying genetic variations. In addition, the results of pQTL and eQTL overlap assessment showed that these protein expression loci may affect the occurrence and development of AA by regulating mRNA transcription levels. Pathway enrichment analysis revealed that proteins associated with AAA were enriched in pathways such as stress response and multicellular biological process regulation, while proteins associated with TAA were mainly involved in biological processes such as cell chemotaxis and immune system regulation. KEGG analysis further showed that these proteins were generally enriched in signaling pathways such as complement-coagulation cascade reaction and cytokine-cytokine receptor interaction. In addition, the drug target evaluation results suggested that among the 450 causally AA-related proteins, nearly two-thirds were classified as druggable or potentially druggable target genes, indicating that intervening these proteins could help develop new drugs for treating AA.

The hundreds of AA-related proteins identified in this study covered multiple key aspects in the pathogenesis and progression of AA, such as extracellular matrix(ECM) remodeling and degradation, inflammation and immunity, oxidative stress, angiogenesis, and vascular remodeling, providing important clues for comprehensively understanding the pathogenesis of this complex disease. Most of these proteins have important biological functions and are potential new targets for the diagnosis and treatment of AA. First, the MR analysis results showed that multiple proteins related to extracellular matrix (ECM) remodeling were significantly associated with AA. These proteins included various matrix metalloproteinases (MMPs), cathepsins, and other ECM-related proteins, such as MMP3, MMP7, MMP8, MMP9, MMP13, CTSB, CTSD, etc. MMPs play a key role in the pathogenesis of AA[16], among which MMP9 is the most extensively studied MMP. It can degrade various extracellular matrix proteins, including elastin and collagen. Studies have shown that MMP9 expression is significantly increased in human AAA tissues, and its plasma levels positively correlate with aneurysm size and expansion rate[17]. CTSB and CTSD (cathepsin B and D) are lysosomal proteases that play a role in extracellular matrix degradation. These proteases are upregulated in AA tissues and may promote aneurysm progression by degrading collagenⅠ[18]. Our drug target analysis showed that multiple ECM-related proteins are targets of existing or potential drugs. For example, MMP9 is a drug target in clinical trials, suggesting that therapeutic strategies targeting the ECM remodeling process may be an effective direction for AA treatment.

Second, our results showed several inflammation-related proteins, such as IL1B, IL2RB, IL5RA, IL17RA, and STAT3, were significantly associated with AA. This is consistent with previous findings. For instance, IL1B (interleukin 1β) is an important pro-inflammatory cytokine, and downregulating IL-1β with antibodies or knocking down IL-1R can inhibit AA progression[19, 20]. STAT3 (signal transducer and activator of transcription 3) is a key regulator of inflammatory response, and blocking downstream inflammatory pathways such as Jak2-Stat3 and IL-6/Jak2/Stat3 signaling can also alleviate AA progression[21, 22]. Our GO enrichment analysis results further supported this view, showing that AA-related proteins were enriched in multiple inflammation and immunity-related biological processes, such as cell chemotaxis and immune system regulation. KEGG pathway analysis also showed that these proteins were enriched in IL-17 signaling pathway and cytokine-cytokine receptor interaction pathway. These findings highlight the central role of inflammation and immune response in the pathogenesis of AA. Our drug target analysis showed that multiple inflammation-related proteins are existing or potential drug targets. For example, IL1B is a successful drug target, providing an important basis for developing anti-inflammatory therapeutic strategies for AA.

Third, our results showed that multiple oxidative stress-related proteins, such as CAT (catalase) and GPX7 (glutathione peroxidase 7), were significantly associated with AA. CAT is an important antioxidant enzyme, and its activity is significantly reduced in AA tissues, which may lead to increased oxidative stress and thus promote aneurysm development[23]. GPX7 is another antioxidant enzyme, mainly involved in lipid peroxidation clearance. Although the role of GPX7 in AA has not been widely studied, there is evidence that oxidative stress plays an important role in the pathogenesis of AA[24]. Oxidative stress may promote AA development through multiple mechanisms, including inducing vascular smooth muscle cell apoptosis, activating MMPs, and promoting inflammatory response. This study highlights the importance of antioxidant mechanisms in maintaining vascular wall stability and suggests potential therapeutic targets. Fourth, this study identified multiple proteins related to angiogenesis and vascular remodeling, such as VEGFR3 (FLT4) and PGF, which were significantly associated with AA. VEGFR3(vascular endothelial growth factor receptor 3)mainly involved in lymphangiogenesis, but also plays an important role in angiogenesis. Studies have found that VEGFR3 expression is increased in AA tissues and may participate in disease progression by promoting neovascularization[25]. Angiogenesis and remodeling may influence AA development through multiple mechanisms, including promoting inflammatory cell infiltration and increasing oxygen and nutrient supply to the vascular wall, thereby supporting pathological remodeling processes. Our drug target analysis showed that some angiogenesis-related proteins are potential drug targets, providing new ideas for developing anti-angiogenic therapeutic strategies for AA.

Fifth, MR analysis results showed that multiple proteins involved in cell signaling transduction, such as AKT2, MAPK13, and MET, were significantly associated with AA. AKT2 is a key member of the PI3K/AKT signaling pathway, involved in cell survival and metabolism regulation. Studies have found that abnormal activation of the AKT signaling pathway is associated with AA formation, and inhibiting AKT can slow aneurysm progression[25]. MAPK13 is a member of the p38 MAPK family, involved in inflammatory response and cellular stress response. Although the specific role of MAPK13 in AA is not yet clear, the p38 MAPK signaling pathway has been shown to play an important role in aneurysm development[27]. These signaling pathway proteins may influence AA development by regulating processes such as cell proliferation, apoptosis, and inflammatory response. Our enrichment analysis results also supported this view, showing that AA-related proteins were enriched in multiple signaling transduction-related biological processes. Sixth, our study also identified multiple proteins involved in metabolic processes, such as ADH1B, ADH4, and ALDH1A1, which were significantly associated with AA. This finding highlights the potential role of metabolic abnormalities in AA pathogenesis, which is a relatively new research direction. Metabolism-related proteins may indirectly participate in AA development by influencing oxidative stress levels, energy metabolism, lipid metabolism, and other processes. Our KEGG pathway analysis also showed that AA-related proteins were enriched in metabolic pathways, further supporting this view. The AA-related proteins in this study are expected to greatly expand the options for AA drug development. Considering the lack of effective drug therapies for AA, this has important implications for advancing the development of precision medicine for AA.

However, some proteins confirmed by previous studies to be associated with AA were not included in this MR analysis, or some of these proteins have not been reported in previous relevant studies, or previous studies have shown that some of these proteins are not related to the pathological mechanisms of AA. For example, some known AA markers, such as serum C-reactive protein (CRP) and D-dimer, have been demonstrated to be closely related to AA and can be used as predictors of aneurysm rupture[28, 29], but were not included in this MR analysis, suggesting that CRP and D-dimer may only be markers of AA rather than causes, or it may be due to differences in research methods and populations. In addition, some factors proven to be involved in AA pathogenesis in animal models, such as Ang-II, suggesting the important role of the Ang-II-AT1R pathway in AA pathogenesis, were not included in this MR analysis, possibly because changes in circulating Ang-II levels do not fully reflect its effects in the local aortic wall. Furthermore, some proteins in the MR analysis, such as COCH, RRM1, CA1, CA6, ITGB7, LAMA4, METAP1, PON3, etc., have not been clearly reported to be associated with AA, and these newly discovered proteins are expected to become research hotspots in the field of AA, providing new entry points for exploring the pathogenesis of AA, but should also be treated with caution, as the causal relationship between these proteins and AA still requires more experimental evidence support. Notably, some proteins considered to be involved in AA pathogenesis did not show a clear causal relationship in this study. For example, MMP2 is upregulated in AA tissues[28, 29], but was not included inthis MR analysis, suggesting that changes in MMP2 expression may be secondary changes rather than initiating causes in the AA development process, and the possibility that limited circulating MMP2 cannot fully reflect local aortic wall MMP2 levels cannot be ruled out. Finally, some proteins in the MR analysis of circulating plasma proteins, such as CA1 and CA6 (carbonic anhydrases 1 and 6, involved in acid-base balance regulation in the kidney), seem to have no obvious relevance to the pathogenesis of AA, and there are currently no reports on the relationship between these proteins and AA, but their presence suggests that they may participate in AA pathogenesis through some unknown mechanisms and have value for further exploration.

This study utilized large-sample proteomics GWAS data from multiple cohorts, constructing the largest MR study on AA to date. The sample size and extensive protein spectrum coverage provided a good data basis for comprehensively evaluating the relationship between circulating proteins and AA, enhancing the reliability and representativeness of the research results. Compared with previous observational studies, MR studies use genetic variations associated with specific protein levels as IVs, based on the principle of random allocation of alleles in Mendel’s law, simulating a process similar to a randomized controlled trial, and can therefore largely avoid confounding bias and reverse causality in traditional epidemiological studies[31]. In addition to the use of standard cis-pQTLs as IVs, trans-pQTLs were also incorporated, capturing more genetic variation and improving the accuracy of causal effect estimates. Multiple MR models and sensitivity analyses, such as IVW, MR-Egger, and weighted median methods, were employed to validate the robustness of the MR results. To further verify the reliability of the associations obtained from the MR analysis, this study also conducted additional analyses, including the Steiger directionality test, colocalization analysis, and pQTL and eQTL overlap assessment. For proteins significantly associated with AA in the MR analysis, this study further conducted pathway enrichment and drug target annotation, which aided in interpreting the biological significance of the MR results from a functional and clinical translational perspective, providing important clues for elucidating the mechanisms of key proteins and developing new diagnostic and therapeutic strategies. However, these bioinformatics analyses are mainly based on existing databases and literature mining results and still require further experimental studies for validation and extension. In addition, due to the complexity of AA pathogenesis, the causal effects of individual proteins may be limited, and the interactions and synergistic effects between proteins may be more important. Future studies need to conduct more network analysis and systems biology research to reveal the regulatory networks and synergistic patterns of proteins in the AA pathogenesis process.

This study also has some limitations. First, the levels and functions of circulating plasma proteins may differ from those of local aortic tissue proteins[6], and this study was unable to directly obtain aortic tissue samples for proteomic testing. This limits the interpretation of the role of local aortic proteomic changes in AA development to some extent. Second, although the population included in this study was mainly European, reducing the impact of population heterogeneity to some extent, the genetic background and protein expression profiles of different races and regional populations may differ, which may affect the generalizability of the MR analysis results to other populations. Third, colocalization analysis is based on the assumption that there is only one shared potential causal variation in a genomic region for each of the two traits[32], while in reality, some regions may have multiple independent causal loci. This may lead to an overestimation of the possibility that some positive MR results are caused by linkage disequilibrium. In the future, more complex methods such as multivariable MR can be attempted to explore the causal structure within specific regions[33]. Fourth, circular RNAs and other non-coding RNAs regulate protein expression at the post-transcriptional level, and this study was unable to incorporate relevant data. Future integration of transcriptomic and proteomic data will help to more comprehensively explain the observed associations. Fifth, this study mainly relied on existing public data information and may not be comprehensive enough in incorporating some of the latest proteomic data. As related research continues to deepen, updating and supplementing the results of this study with the latest protein-related association data will be a valuable development direction.

In summary, this study used large-scale plasma proteomic association data and a MR analysis framework to systematically evaluate the causal relationship between circulating protein levels and AA at the wproteome-wide level, identifying a large number of potentially new pathogenic proteins and therapeutic targets for AA, providing important references for elucidating the molecular pathogenesis of AA and advancing drug development.

## Data Availability

All data produced are available online at the website https://www.finngen.fi/en

